# Photon-Counting Computed Tomography of Degradable Bone Cement Loaded with Gadolinium Nanoparticles

**DOI:** 10.1101/2025.10.23.25338635

**Authors:** Anna Marks, Kitz Paul D. Marco, Allan John R. Barcena, Marvin R. Bernardino, Megan C. Jacobsen, Rick R. Layman, Marites P. Melancon

## Abstract

**Objectives:** Polymethylmethacrylate (PMMA)-based cements cannot integrate with endogenous bone over time, leading to long term instability and risk of subsequent fractures. This has led to the development of absorbable bone cements that address many of the limitations of PMMA. Often, visualization of absorbable bone cements is challenging because their radiopacity closely resembles that of vertebral bone. This study aims to improve the radiopacity of absorbable bone cements through the addition of gadolinium nanoparticles (GdNP). In addition, we aim to determine whether photon counting computed tomography (PCCT), which has been shown to have enhanced image quality when compared with conventional CT, provides better contrast ratio between GdNP-loaded bone cement and vertebral bone than energy-integrating detector CT (EID-CT). Finally, we aim to evaluate if PCCT can successfully quantify the concentration of GdNPs through imaging.

**Materials and Methods:** GdNPs were synthesized using a one-pot thermal decomposition method and characterized using transmission electron microscopy and dynamic light scattering. Hydroxyapatite-based bone cement was loaded with varying concentrations of GdNPs (0-10 % w/w), and the contrast ratio between the cement and vertebral bone was evaluated using preclinical EID-CT and PCCT scanners. Gadolinium standards (0-20 mg/mL) were imaged using a preclinical PCCT. Attenuation of the cement was measured in each PCCT energy bin, and the concentration of the gadolinium was estimated using gadolinium material decomposition images.

**Results:** The synthesized GdNPs had a mean diameter of 15.42 ± 1.82 nm. Signal intensity increased with increasing concentration of GdNPs for both EID-CT and PCCT. In EID-CT images, the 8% and 10% GdNP-loaded bone cement had higher contrast ratios relative to bone than the conventional bone cement (p < 0.01). The contrast ratio between the 10% GdNP-loaded bone cement significantly differed from that of conventional bone cement for all PCCT energy bins (p < 0.05). The 42-51 keV energy bin yielded the most significant difference between 10% GdNP-loaded bone cement and bone cement with no GdNP loading. Contrast ratios obtained from PCCT images differed significantly from EID-CT contrast ratios (p < 0.0001). Estimated gadolinium concentrations were highly correlated with true gadolinium concentrations (r = 0.999). PCCT was able to accurately quantify gadolinium concentrations with a root mean square error of 1.60 mg/mL.

**Conclusions:** The use of GdNPs led to a higher cement-vertebra contrast ratio for both EID-CT and PCCT. Overall, PCCT demonstrated higher contrast ratios than EID-CT. Material decomposition successfully quantified the amount of gadolinium present in samples and allowed for improved visual differentiation of the GdNP**-**loaded bone cement from the calcium-based vertebral bodies. Thus, the incorporation of radiopaque GdNPs and imaging with PCCT improved visualization of the bone cement and enabled GdNP quantification, which may lead to improved implant monitoring.

## 1. Introduction

Percutaneous vertebroplasty and kyphoplasty are minimally invasive procedures commonly used to treat vertebral compression and pathological fractures. Vertebral compression fractures are commonly experienced by patients with osteoporosis, while pathological vertebral fractures are commonly caused by osseous tumor involvement.^1^ Polymethylmethacrylate (PMMA) is the most widely used bone cement for these procedures because it is easy to use, cost effective, and has considerable mechanical strength.^2^ However, PMMA cannot integrate with endogenous bone, which can affect the long-term stability of the bone implant. Furthermore, PMMA has very different mechanical properties compared to the surrounding bone, which can cause adjacent vertebral fractures.^2^ Degradable bone cements, such as calcium phosphate- and hydroxyapatite (HA)-based cements, have been developed to address these limitations. HA supports bone formation, which can improve implant stability when compared with PMMA. In addition, HA can be used to create bioabsorbable cements that allow for the load to be slowly transferred from the implant to the bone. This property can induce bone growth and potentially decrease the risk of complications after implantation.^3^

Radiopacity is important for bone cements to allow adequate visualization during the procedure and in follow-up scans. Cements without contrast agents have a slightly higher CT number than bone at implantation; however, post-procedural monitoring of the implant can be difficult as the material degrades.^4^ To address this limitation, many contrast agents have been integrated into bone cement materials to improve visualization on computed tomography (CT) imaging.^4–7^ Specifically, gadolinium nanoparticles (GdNP) have shown promise for increasing the contrast of bone cement for both CT and magnetic resonance (MR) imaging.^8^ Gadolinium (Z = 64), traditionally an MR contrast agent, has a higher atomic number than iodine (Z = 53), which is widely used as a CT contrast agent. Due to gadolinium’s higher atomic number, it can efficiently attenuate X-rays, leading to higher image contrast when compared to the same concentration of iodine.^9^ However, while gadolinium-based contrast agents are considered to be safe in lower doses, exposure to high amounts of free gadolinium has been associated with nephrogenic systemic fibrosis, a serious renal disease.^10^ GdNPs have demonstrated very low release of free gadolinium in the body which reduces toxicity when compared to traditional gadolinium-based contrast agents.^11,12^ This makes GdNPs a safer alternative than traditional gadolinium contrast agents, and their high atomic number and electron density make them promising candidates as CT contrast agents.

Photon counting CT (PCCT) is a recent computed tomography technology that may also improve the ability to distinguish NP-infused bone cement from vertebral bone *in vivo*. PCCT detectors employ a semiconductor-based detector that allows each x-ray photon to be directly converted into an electrical signal proportional to photon energy, unlike energy integrating detectors (EID), which require photons to be converted to visible light before a signal can be produced. As a result, every element in a PCCT detector measures the energy of each incident photon, allowing for effective removal of electronic noise and the sorting of each signal into prespecified energy bins. Compared to EID-CT, PCCT images have lower noise, increased contrast-to-noise ratio (CNR), and may be configured to identify and quantify materials based on their x-ray K-edge.^13^ Since the PCCT can sort incident photons into corresponding energy bins, PCCT can provide improved material decomposition when compared to dual energy CT (DECT),particularly for materials that have k-edges within the diagnostic energy range such as gadolinium (k-edge = 50.2 keV).^14,15^ On systems with selectable energy bins, the bins can be chosen such that one is below the k-edge energy, and another is above it to increase image contrast and allow for improved material decomposition. Halttunen et al.^16^ evaluated the performance of GdNPs as a contrast agent for PCCT *in vivo* and found that GdNPs were a good candidate for use in PCCT. In addition, PCCT is able to estimate the concentrations of material present in images, and it has been shown that PCCT can accurately estimate the concentration of gadolinium *in vivo*.^17,18^

In this paper, we incorporated GdNPs into absorbable bone cement and evaluated whether PCCT could improve the visualization of bone-implant interfaces when compared to EID-CT. We hypothesize that the addition of GdNP-based contrast agents will further improve the ability of HA-based bone cement to be distinguished from vertebral bone.^19^ Additionally, in this proof-of-concept, preclinical study, we show that PCCT may allow for Gd concentration to be reliably quantified.

## 2. Methods

### 2.1 Materials

Gd acetate, oleic acid, oleylamine, dichloromethane, and hydroxyapatite were obtained from Sigma-Aldrich (St. Louis, MO, USA). Gadolinium standard solution (10,021 ± 35 μg/mL) in 7% v/v HNO_3_ was obtained from Inorganic Ventures (Christianburg, VA, USA) and used to prepare standard Gd solutions of varying concentrations from 2 to 20 mg/mL.

### 2.2 GdNP Synthesis and Characterization

GdNPs were synthesized in a 500 mL three-neck flask. First, 1.67 g of Gd acetate (5 mmol) was added to 30 mL of oleic acid (90 mmol) and 100 mL of oleylamine (300 mmol). The mixture was heated to 120 °C for 1 hour under an argon atmosphere while magnetically stirred. After 1 hour, the temperature was increased to 280 °C and maintained for 6 hours. The solution was then allowed to cool at room temperature. After cooling, the solution was washed three times with a 1:1 solution of ethanol:acetone followed by centrifugation. The precipitate was redispersed in dichloromethane (DCM) and stored until further use.

The size and distribution of the GdNPs were visualized using transmission electron microscopy (TEM) imaging with a JEM 1010 microscope (JEOL USA, Peabody, MA, USA) and the size and distribution were quantified using ImageJ software, version 2.16.0 (U. S. National Institutes of Health, Bethesda, Maryland, USA). Electrophoretic potential was determined using dynamic light scattering (NanoBrook 90Plus PALS, Brookhaven).

### 2.3 Fabrication of GdNP Loaded Bone Cement

Bone cement was created using a HA base with the addition of varying concentrations (0%, 2%, 4%, 6%, 8%, 10% w/w) of GdNPs dispersed in DCM. The resulting powder was then heated for 4 hours at 40.5 °C to allow for the evaporation of DCM. After heating, citric acid buffer (50 mM, pH 6.5) was added to the powder to create a paste, which was placed in molds and allowed to dry into disks for imaging.

### 2.3 Preparation of Agar-Based Vertebral Phantom Using Rat Vertebrae

Rat lumbar vertebrae were harvested and cleaned following a protocol approved by MD Anderson’s Institutional Animal Care and Use Committee (00001882-RN02). A 1.2 mm diameter × 2 mm depth cylindrical hole was drilled on the anterior surface of the vertebral body and filled with the HA-based bone cement prepared as described previously. The resulting cement-filled vertebrae were placed into a phantom made of 1.5% w/v agarose in water (Supplementary Figure 1).

### 2.4 EID-CT and PCCT acquisition and image analysis

The phantom was imaged using two small animal CT systems: EID-CT micro-CT (Skyscan 1276; Bruker, Billerica, MA) and PCCT micro-CT (Microlab 5×120; MARS Bioimaging Ltd, Christchurch, New Zealand). EID micro-CT was performed at 70 kVp tube voltage, 200 µA tube current, and 135 ms rotation time. EID-CT image reconstruction was performed using filtered back projection with a slice thickness of 0.04 mm and pixel size of 0.04 x 0.04 mm^2^.

PCCT was performed at 118 kVp, 40 µA, and 160 ms rotation time. The energy bins were optimized to generate approximately equal counts in each bin while providing bins close to the k-edge for gadolinium (7-30, 30-42, 42-51, 51-59, 59-118 keV). PCCT image reconstruction was performed using the MARS proprietary algebraic reconstruction algorithm to generate bin images and material decomposition images for the following basis materials: water, adipose, calcium, gadolinium, iodine, and gold. Images were reconstructed with a slice thickness of 0.09 mm and pixel size of 0.09 x 0.09 mm^2^. Of note, the energy bin images are generated in units of linear attenuation coefficient (µ [cm^−1^]) rather than Hounsfield units (HU). For direct comparison of the two modalities, we converted µ to HU using the following formula:

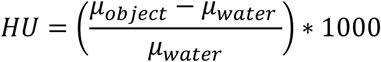

Where µ_water_ was measured from a PCCT scan of deionized water for each energy bin (Supplementary Table 1).

The contrast ratio of GdNP-loaded bone cement to vertebral bone was calculated from each of the EID-CT images for each GdNP concentration. HU values were measured for GdNP-loaded bone cement and vertebral bone using regions of interest with an area greater than 100 pixels^2^ (Supplementary Figure 2). Contrast ratio was calculated using the following equation:

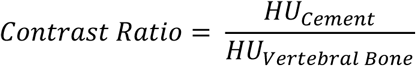

From PCCT images, the contrast ratio between bone cement and vertebral bone were calculated for every energy bin and GdNP concentration. In addition to the phantom, five 1.5 mL vials of gadolinium standards at varying concentrations (2, 5, 10, 15, 20 mg/mL) were prepared from gadolinium standard solution (10,021 ± 35 μg/mL) in 7% v/v HNO_3_ and inserted into a cylindrical PMMA phantom with a 100 nm diameter provided by MARS Bioimaging. Standards were imaged on the PCCT system and the estimated concentration of Gd was measured on the gadolinium-specific image and compared to the nominal concentration of the standards. Root mean square error was calculated to evaluate the accuracy of the concentration estimation.

### 2.4 Statistical Analysis

All studies were conducted in triplicate (n=3). Analysis was performed using GraphPad Prism, version 10.3.1 software (GraphPad, San Diego, CA, USA). All data is presented as mean ± standard deviation. The Shapiro-Wilk test was used to test for normality. One-way analysis of variance (ANOVA) and Dunnett’s multiple comparison test, using 0% gadolinium HA bone cement as the control sample, was used to compare the differences in contrast ratio between each GdNP concentration and the control. Two-way ANOVA and Holm-Sidak multiple comparisons test was used to evaluate differences between the contrast ratios calculated from EID-CT and PCCT at varying GdNP concentrations. Pearson correlation was used to determine the relationship between estimated and true gadolinium concentration. For all statistical tests, p < 0.05 was considered to be statistically significant.

## 3. Results

### 3.1 GdNP Synthesis and Characterization

The chemical synthesis successfully yielded GdNPs with a uniform size and shape distribution as seen in the TEM images shown in Figure 1A-B. Resulting GdNPs were spherical shape with a mean diameter of 5.42 ± 1.85 nm. The electrophoretic potential of the GdNPs was -15.81 ± 1.45 mV.

**Figure 1:**
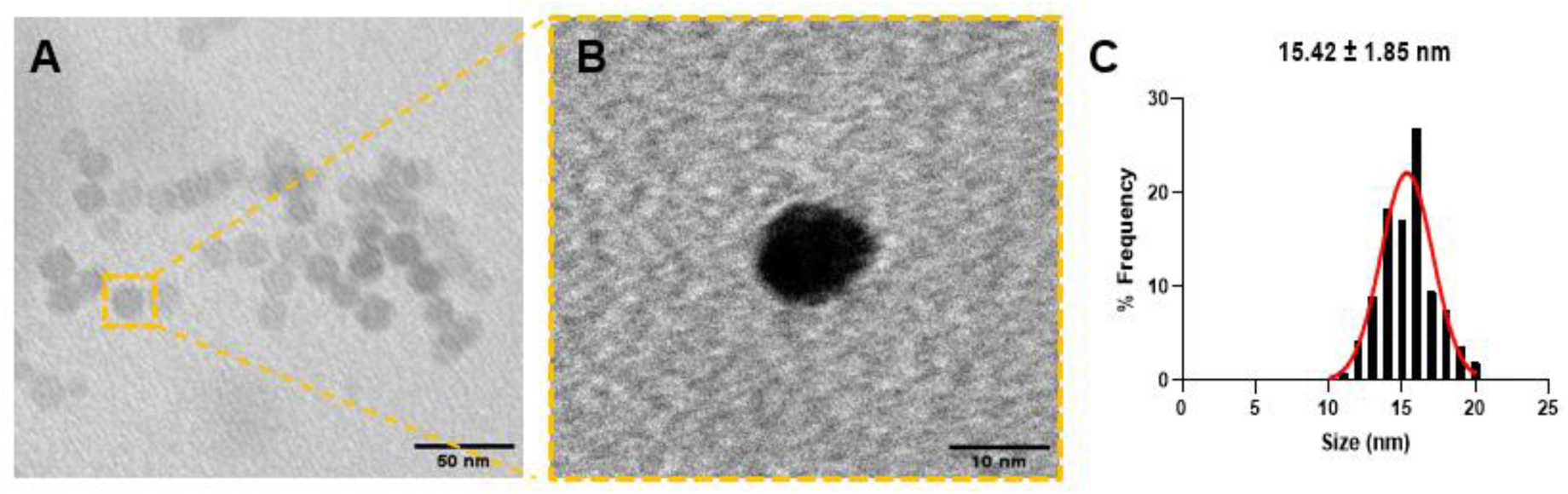
Synthesis and characterization of GdNPs. A. TEM image of GdNPs showing their spherical morphology and uniform size distribution. B. High magnification TEM image highlighting particle morphology and size. C. Size distribution of GdNPs analyzed using ImageJ (v2.16.0), with a mean diameter of 15.42 ± 1.85 nm.

### 3.2 EID-CT and PCCT Imaging

The visible contrast between the GdNP-loaded bone cements increases with increasing GdNP concentration for both EID-CT and PCCT, as shown in Figure 2. In EID-CT images, the difference in contrast ratio was significant for the 8% and 10% w/w GdNP-loaded bone cements when compared to the bone cement with no GdNPs added (p < 0.01) (Figure 3). The contrast ratio increased significantly between the conventional bone cement and the 10% GdNP w/w loaded bone cement on PCCT for all energy bins (p < 0.05) (Figure 4). In the 42-51 keV energy bin, both the addition of 8% GdNP (p < 0.05) and 10% GdNP (p<0.001) had the most significant improvements in contrast ratios compared to cement without GdNP loading (Table 2).

**Figure 2:**
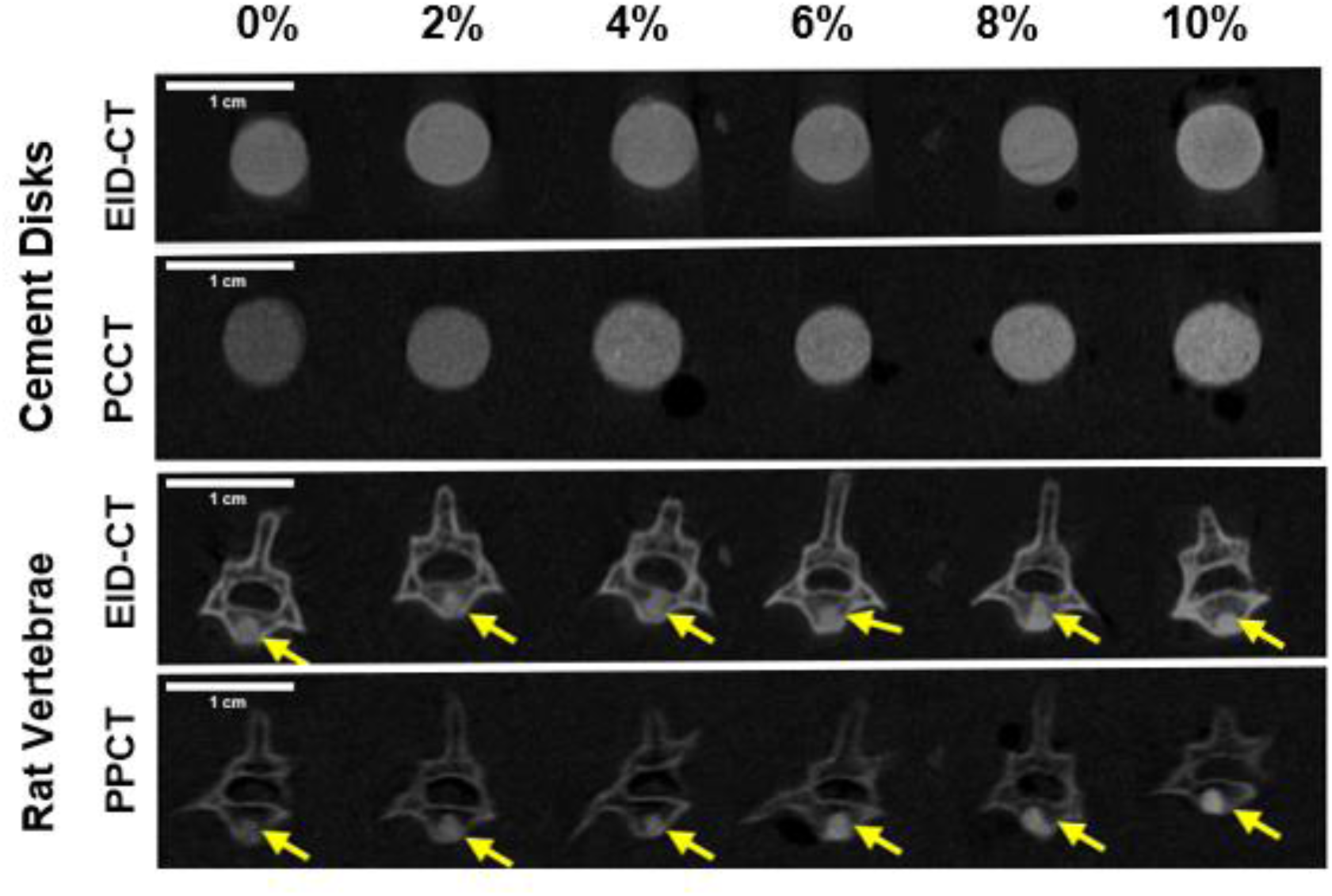
EID-CT (Window/Level: 20341/9593 HU) and PCCT (Window/Level: 1.576 cm^−1^/0.8759 cm^−1^, Energy Bin: 42-51 keV) phantom images of bone cement disks and cements in spine. From top to bottom: EID-CT images of GdNP-loaded bone cement disks; PCCT images of GdNP-loaded bone cement disks; EID-CT images of vertebral bone filled with GdNP-loaded bone cement; PCCT images of vertebral bone filled with GdNP-loaded bone cement. For both EID-CT and PCCT, cement signal increases with increasing GdNP concentration. PCCT shows greater visible contrast between the vertebrae and the cement (yellow arrow).

**Figure 3:**
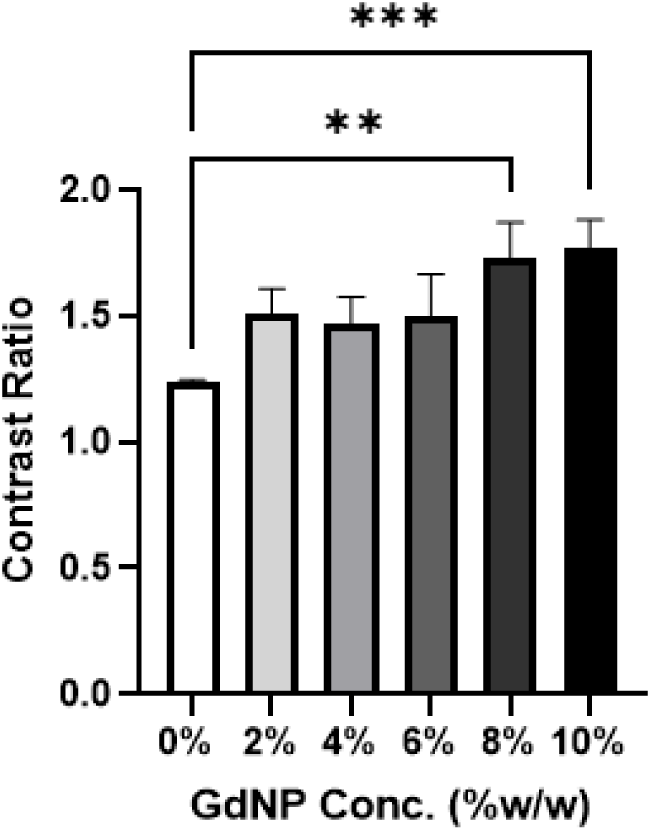
Contrast ratio of GdNP-loaded bone cement to vertebral bone for EID-CT images. Bone cement loaded with 8% and 10% w/w GdNP had a significantly greater contrast ratio than the bone cement with no GdNP loading. *p < 0.01 **, p< 0.001 \*\*\**.

**Figure 4:**
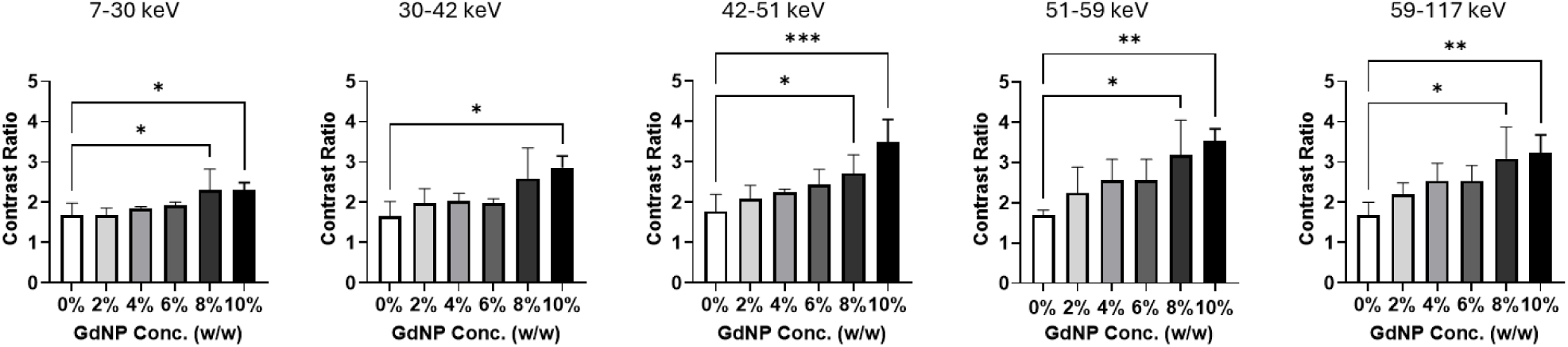
Contrast ratio of GdNP-loaded bone cement to vertebral bone for each PCCT energy bin. The energy bin from 42-51 keV yielded the greatest amount of differentiation between GdNP cement groups. Bone cement loaded with 10% w/w GdNP had a significantly greater contrast ratio than the bone cement with no GdNP loading for all energy bins. *p < 0.05 *, p 0.01 **, p < 0.001 \*\*\**.

When quantitatively compared to EID-CT, the use of PCCT yielded statistically significant contrast ratios for both the 8 and 10% w/w GdNP-loaded bone cements at every energy bin (p < 0.05). Two-way ANOVA determined that there was a statistically significant difference between the contrast ratios from the EID-CT images and the contrast ratios from the PCCT images (p < 0.0001) for all energy bins. The difference between EID-CT and PCCT is particularly apparent in the 42-51 keV, 51-59 keV, and 59-117 keV energy bins, where significant improvement in contrast ratio was seen in as low as 4% w/w GdNP. The 42-51 keV energy bin showed the most significant difference between the EID-CT contrast ratio and PCCT contrast ratio for the 10% w/w GdNP-loaded bone cement (p < 0.0001) (Table 1, Figure 5).

**Figure 5:**
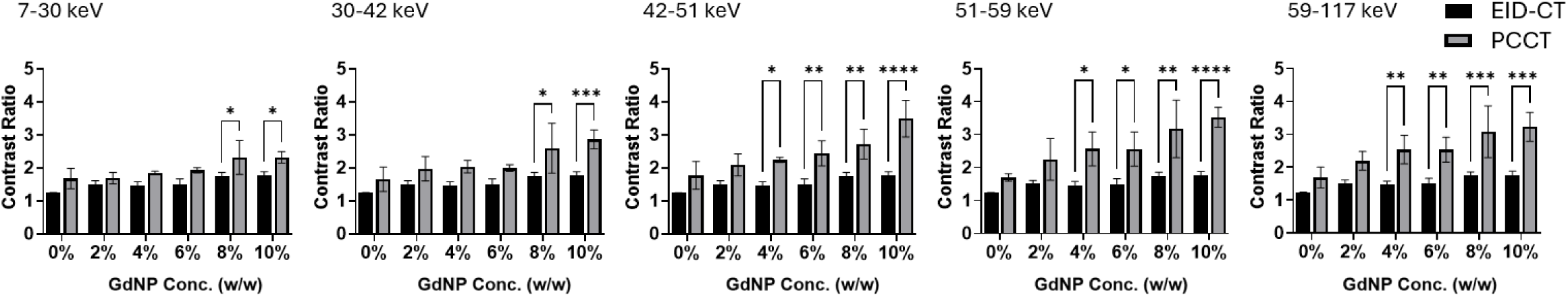
Comparison of resulting contrast ratios from EID-CT and each PCCT energy bin. There was a statistically significant difference between EID-CT and PCCT contrast ratios for all PCCT energy bins for 8 and 10% w/w GdNP. The 42-51 keV energy bin had the most significant difference between EID-CT and PCCT contrast ratio for the 10% w/w GdNP-loaded cement. *p < 0.05 *, p < 0.01 **, p < 0.001 ***, p < 0.0001 \*\*\*\**.

**Table 1:** Numerical comparison of resulting contrast ratios for EID-CT and each PCCT energy bin.

|  | **EID-CT** | **PCCT** | **PCCT** | **PCCT** | **PCCT** | **PCCT** |
| --- | --- | --- | --- | --- | --- | --- |
|  |  | **7- 30 keV** | **30-42 keV** | **42-51 keV** | **51 -59 keV** | **59 -117 keV** |
| **0%**<br>**w/w**<br>**GdNP** | 1.24 ($\pm 0.01$) | 1.68 ($\pm 0.25$) | 1.65 ($\pm 0.30$) | 1.77 ($\pm 0.34$) | 1.70 ($\pm 0.09$) | 1.69 ($\pm 0.26$) |
| **2%**<br>**w/w**<br>**GdNP** | 1.51 ($\pm 0.08$) | 1.69 ($\pm 0.14$) | 1.97 ($\pm 0.30$) | 2.09 ($\pm 0.27$) | 2.25 ($\pm 0.52$) | 2.20 ($\pm 0.23$) |
| **4%**<br>**w/w**<br>**GdNP** | 1.47 ($\pm 0.09$) | 1.85 ($\pm 0.04$) | 2.03 ($\pm 0.15$) | 2.24 ($\pm 0.07$) | 2.57 ($\pm 0.42$) | 2.54 ($\pm 0.35$) |
| **6%**<br>**w/w**<br>**GdNP** | 1.50 (± 0.14) | 1.93 (± 0.07) | 1.99 (± 0.08) | 2.44 (± 0.31) | 2.56 (± 0.42) | 2.54 (± 0.31) |
| **8%**<br>**w/w**<br>**GdNP** | 1.73 (± 0.12) | 2.31 (± 0.42) | 2.59 (± 0.62) | 2.72 (± 0.37) | 3.18 (± 0.71) | 3.08 (± 0.64) |
| **10%**<br>**w/w**<br>**GdNP** | 1.77 (± 0.09) | 2.31 (± 0.14) | 2.86 (± 0.24) | 3.49 (± 0.45) | 3.53 (± 0.25) | 3.23 (± 0.36) |

**Table 2:** Mean estimated concentration (mg/mL) for each standard concentration (mg/mL). The root mean square error in estimated gadolinium concentration was 1.60 mg/mL.

| Standard<br>Concentration<br>(mg/mL) | Mean Estimated<br>Concentration (mg/mL) |
| --- | --- |
| 2 | 3.37 (± 0.208) |
| 5 | 6.88 (± 0.456) |
| 10 | 11.5 (± 0.671) |
| 15 | 16.4 (± 1.15) |
| 20 | 21.3 (± 0.511) |

There was an excellent correlation between estimated and true gadolinium concentration values (r = 0.999, p < 0.00001). The root mean square error in gadolinium quantification was 1.60 mg/mL (Table 2). The maximum difference between the estimated and standard gadolinium concentration was 2.55 mg/mL. Performance of gadolinium concentration estimations by PCCT material decomposition is shown in Figure 6. CT number increased with increasing gadolinium concentration for all energy bins. Material decomposition images were also able to improve the visualization of GdNP loaded bone cement as shown in Figure 7. The 10% w/w GdNP-loaded bone cement showed the best delineation between bone cement and vertebral bone. At least 4% w/w GdNP was needed to visualize the cement in the vertebrae using material decomposition. The 2% w/w GdNP-loaded bone cement was not identified solely as gadolinium by the material decomposition due to the implant’s calcium content and therefore, could not be visualized using the material decomposition images.

**Figure 6:**
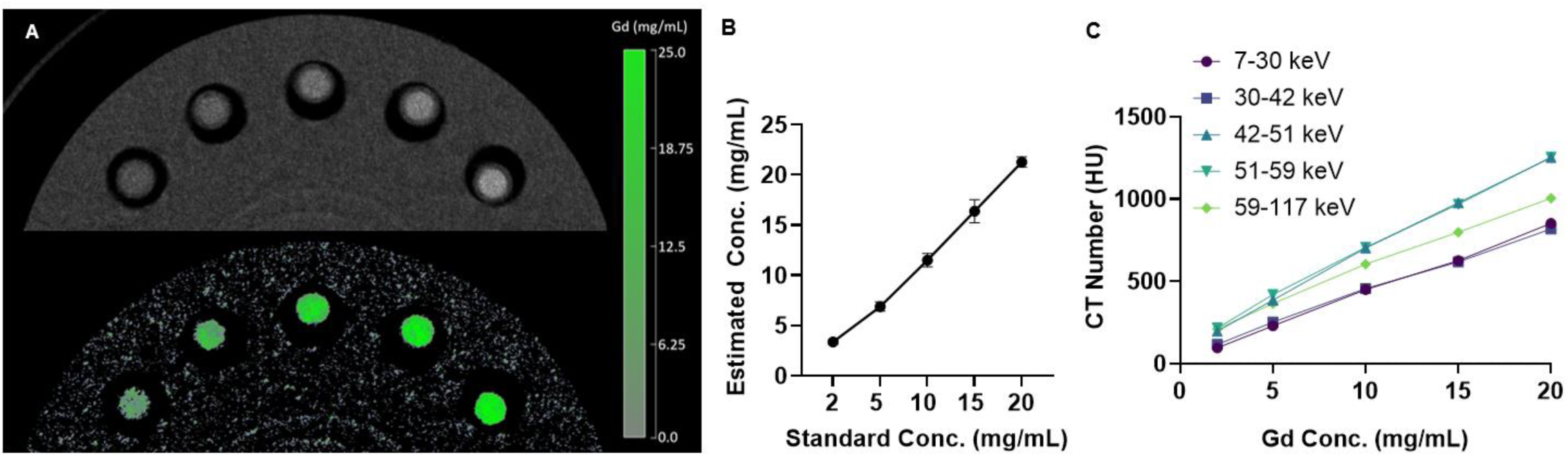
A. PCCT images of gadolinium standards and the corresponding material decomposition. From left to right: 2, 5,10,15, 20 mg/mL gadolinium. Material decomposition images show increasing ability to recognize gadolinium (represented with green) as standard gadolinium concentration increases. B. Estimated concentration of gadolinium measured from PCCT material decomposition plotted against the standard gadolinium concentration. C. CT number plotted against standard gadolinium concentration for each energy bin. Material decomposition allowed for accurate quantification of gadolinium standards with a root mean square error of 1.60 mg/mL. CT number scaled linearly with gadolinium concentration for each energy bin.

**Figure 7:**
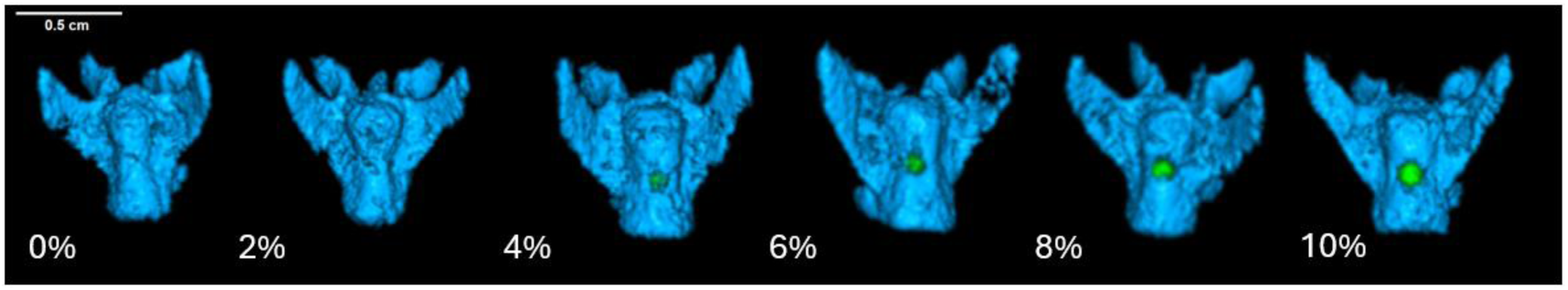
PCCT material decomposition images of GdNP-loaded bone cement implanted in vertebral spines. Gadolinium bin is denoted with green, and calcium bin is denoted with blue. As GdNP concentration increased in the bone cement, PCCT material decomposition showed improved ability to distinguish GdNP-loaded cement from vertebral bone.

## 4. Discussion

In this study, we successfully synthesized uniform GdNPs with an average diameter of 15 nm and a negative electrophoretic potential, both characteristics that suggest good colloidal stability and suitability for incorporation into bone cement.^20^ When incorporated into HA-based cement, GdNPs provided concentration-dependent increases in radiographic visibility across both EID-CT and PCCT. These results confirm our initial expectation that higher GdNP content would enhance contrast, while also demonstrating the superior performance of PCCT over EID-CT for distinguishing material-specific signals.^16^

In our analysis, PCCT yielded consistently higher contrast ratios than EID-CT, underscoring the advantage of photon-counting systems in discriminating materials with similar attenuation properties.^21^ Significant differences in contrast ratio were observed for both 8% and 10% w/w GdNP-loaded bone cements compared to 0% w/w cements, with 10% w/w cements showing the most robust contrast enhancement across all energy bins. Importantly, the 42–51 keV bin produced the most pronounced differences between groups, which is consistent with the gadolinium K-edge at 50.2 keV.^22^ At this energy, the mass attenuation coefficient of gadolinium diverges sharply from that of calcium, producing a measurable increase in CT number and contrast ratio (Supplementary Figure 3). While we expect the gadolinium attenuation to be significantly higher in the 51-59 keV bin compared to the 42-41 keV bin, it must be noted that there may be significant overlap between the energy bins due to detector effects (i.e. charge sharing, K-escape fluorescence) that distort the energy spectrum. This finding highlights the importance of energy bin selection in PCCT, where careful alignment with elemental K-edges can yield substantial gains in visualization compared to conventional EID-CT, which lacks photon energy discrimination.^14^

The ability of PCCT to perform material decomposition further strengthens its clinical potential. PCCT material decomposition has been shown to be a clinical useful tool in multiple disciplines.^23^ In orthopedics, specifically, PCCT material decomposition has been successfully used to improve bone implant visualization and improve identification of implant failures when compared with conventional imaging modalities.^19,24^ Additionally, PCCT has successfully been used to separate gadolinium from HA and gadolinium from calcium.^21,25^ Similarly to these studies, we found that gadolinium and calcium were accurately separated, allowing bone cement to be distinguished from vertebral bone despite both being highly attenuating materials. Of note, while the 2% gadolinium standard with an aqueous background matrix was readily identified as gadolinium by the material decomposition algorithm, the 2% w/w GdNP-loaded cement was labeled as predominately calcium. In contrast, concentrations ≥ 4% were clearly identified as containing gadolinium. While misclassification by the visualization software occurs at lower gadolinium concentrations, higher Gd loading improves accurate material identification. However, increasing the Gd content may also compromise the cement’s mechanical integrity. Therefore, future work should focus on optimizing formulations that balance imaging performance with mechanical strength. While 10% w/w GdNP cements appear promising for visualization, further investigation is needed to confirm that its compressive strength and handling characteristics remain comparable to clinical standards.^26^ Similarly, the biocompatibility of GdNP-loaded cements must be established, as degradation over time may release nanoparticles that could impact local cell survival or systemic biodistribution.^27^

Quantitative performance of the PCCT was also excellent, with estimated gadolinium concentrations correlating almost perfectly with true values (r = 0.999) and a root mean squared error of 1.60 mg/mL. These levels of error are consistent with the other studies which used PCCT images to quantify gadolinium. For example, Ren et al.^17^ and Tao et al.^18^ reported root mean square errors of 1.00 mg/mL and 2.02 mg/mL, respectively, when using PCCT material decomposition to estimate gadolinium concentration. Our results confirm that PCCT material decomposition can be used to quantify the amount of gadolinium present in a sample and to improve the visualization of two highly attenuating materials (bone and GdNPs).

While the use of PCCT and GdNPs show promise to improve visualization of absorbable bone cement our study had some imaging limitations. For example, the calibration of gadolinium and material decomposition could be further optimized on our pre-clinical PCCT.^28^ Additionally, spatial resolution of the EID-CT used in the study is better than the PCCT. As a result, overall image quality of the images obtained from the PCCT and EID-CT could not be fairly evaluated. Finally, no noise measurements were made since the Gd-loaded bone cement was expected to be non-uniform.

This study demonstrates that incorporating GdNPs into HA-based bone cement significantly enhances radiopacity in both EID-CT and PCCT imaging. Importantly, PCCT provided superior visualization compared to conventional EID-CT, particularly through its ability to perform material decomposition and exploit the gadolinium K-edge for material-specific contrast. These advantages enabled clear differentiation between bone cement and vertebral bone, as well as accurate quantification of gadolinium standards. Together, these findings highlight the potential of GdNP-loaded bone cements not only for improved intra- and post-procedural visualization, but also for noninvasive longitudinal monitoring of implant integrity, degradability, and bone integration. Future studies should extend these findings in vivo, evaluate the mechanical and biological properties of GdNP-loaded cements, and explore longitudinal monitoring of implant degradation. Ultimately, this strategy may provide a powerful tool for improving the radiopacity and traceability of bone cement implants, thereby enhancing long-term patient management.

## Data Availability

All data produced in the present study are available upon reasonable request to the authors.

## Abbreviations

ANOVA: analysis of variance
CNR: contrast to noise ratio
CT: computed tomography
DCM: dichloromethane
DECT: dual-energy computed tomography
EID-CT: energy integrating detector computed tomography
GdNP: gadolinium nanoparticles
HA: hydroxyapatite
MR: magnetic resonance
PCCT: photon counting computed tomography
PMMA: polymethylmethacrylate
TEM: transmission electron microscopy

