## Supplemental Figures for "Photon-Counting Computed Tomography of Degradable Bone Cement Loaded with Gadolinium Nanoparticles"

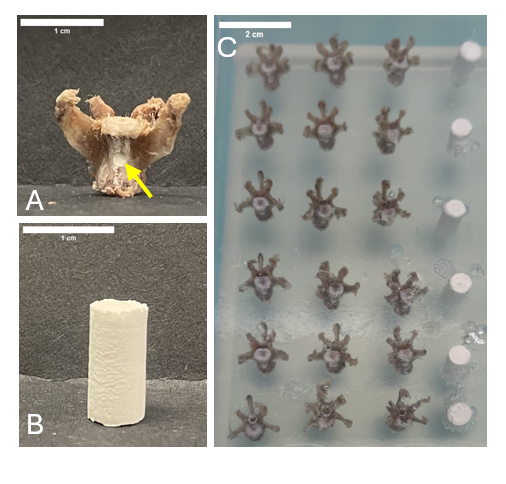
**Supplementary Content:**

**Supplementary Figure 1**: A. Vertebra filled with GdNP-loaded bone cement. B. GdNP- loaded bone cement disk. C. Agar-based vertebral phantom. Three vertebral bodies harvested from rats were filled with GdNP-loaded bone cement for each concentration of gadolinium. GdNP-loaded bone cement disks, shown on the right side of the phantom, corresponding to each concentration were also imaged for material decomposition analysis.

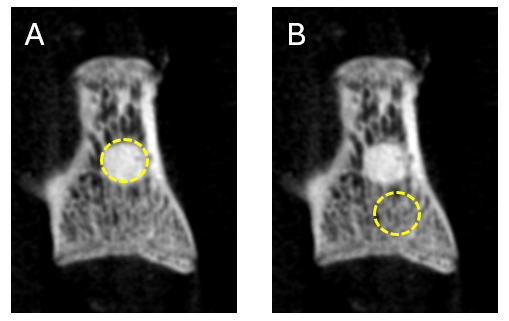

**Supplementary Figure 2:** Examples of the regions of interest (ROI) used to calculate the contrast ratio of bone cement to vertebral bone. A. ROI used to measure the HU value of bone cement. B. ROI used to measure the HU value of vertebral bone.

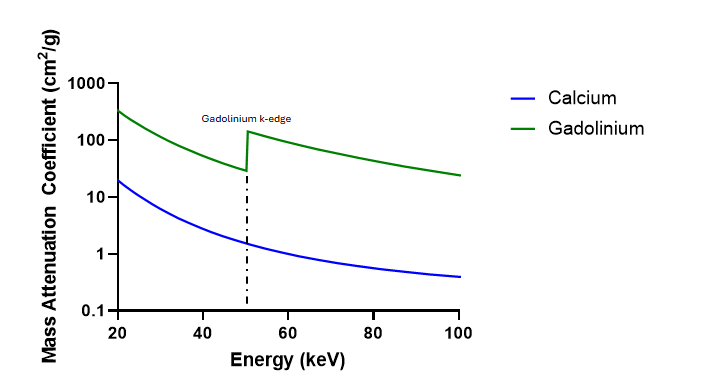

**Supplementary Figure 3:** Mass attenuation coefficients of calcium and gadolinium at energies within the diagnostic region. At the k-edge energy of gadolinium (50.2 keV) the difference in the mass attenuation of calcium and gadolinium is the greatest.

**Supplementary Table 1:** Linear attenuation of water for each energy bin. Values were determined from measurements taken from PCCT images for each energy bin.

| **Energy Bin (keV)** | **µ_water_ (cm^-1^)** |
| --- | --- |
| 7-30 | 0.205 |
| 30-41 | 0.196 |
| 41-52 | 0.171 |
| 52-59 | 0.160 |
| 59-117 | 0.157 |
